# Trait Mindful Nonreactivity and Nonjudgment Prospectively Predict of COVID-19 Health Protective Behaviors Across a Two-Month Interval in a USA Sample

**DOI:** 10.1101/2021.07.21.21260971

**Authors:** William H. O’Brien, Shan Wang, Aniko Viktoria Varga, Chung Xiann Lim, Huanzhen Xu, Somboon Jarukasemthawee, Kullaya Pisitsungkagarn, Piraorn Suvanbenjakule, Abby Braden

## Abstract

The COVID-19 pandemic has prompted a growing recommendation for social distancing and using personal protective equipment (PPE) to help mitigate the virus transmission. Previous studies have shown promising relationships between perceived susceptibility to COVID-19, mindfulness-related variables, and COVID-19 health protective behaviors (social distancing and PPE use). In this longitudinal study, the variables were measured across a two-month interval during the earlier phase of the pandemic in June (Time 1) and August (Time 2), 2020. The results from 151 matched USA MTurk participants indicated that the perceived susceptibility to COVID-19 did not significantly predict the health protective behaviors. For mindfulness, nonreactivity was positively related to PPE use while nonjudgement was negatively related to PPE use. Accordingly, mindfulness promotion messages could be a way to increase the likelihood of people performing health protective behaviors to better constrain the COVID-19 outbreak.

## Introduction

PPE use and social distancing substantially reduce the risk for transmitting and contracting COVID-19 (Anfinrud, et al., 2020; Howard et al., 2020; Leung et al., 2020; Social distancing: Courtemanche, et al., 2020; Wu, et al., 2020). Lyu and Wehby (2020) demonstrated that the implementation of mask mandates in the USA was associated with the prevention of approximately 200,000 new COVID-19 cases across a two-month time span. A meta-analytic review of 172 studies by Chu et al., (2020) demonstrated that PPE use and social distancing were reliably associated with a significant reduction in risk for contracting COVID-19 among the general population and healthcare workers.

Perceived susceptibility is a well-established predictor of health protective behaviors such as diet, safety adherence, attending cancer screenings, and obtaining vaccinations (Brewer, et al., 2007; O’Brien, et al., 1995; Sheeran, et al., 2014). Perceived susceptibility has also been identified as a correlate of PPE use and social distancing in non-COVID-19 contexts (Bish & Michie, 2010). Sim, Moey, and Tan (2014) concluded that perceived susceptibility was the most consistent predictor of facemask use in community settings *prior to COVID-19*. Similarly, Tang and Wong (2004) reported that perceived susceptibility significantly predicted facemask use over and above demographic variables during the SARS outbreak.

Perceived susceptibility to COVID-19 specifically has been associated with social distancing. In a cross-sectional study, de Bruin and Bennett (2020) found that higher perceived susceptibility to COVID-19 infection was significantly correlated with reported social distancing behaviors. Wise and colleagues (2020) also identified a positive cross-sectional association between perceived likelihood of contracting COVID-19 and social distancing and hand washing.

The relationships among health threat stimuli (e.g., public health messaging), perceived susceptibility, and health protective behaviors relevant to COVID-19 can be examined from a functional contextual perspective (Vilardaga, et al., 2009; Zhang, et al. 2017). Exposure to COVID-19 health threat stimuli evokes increased levels of perceived susceptibility and anxiety (Contrada & Baum, 2011; Hyde, et al., 2019). By engaging in health protective behaviors such as PPE use and social distancing, an individual can reduce perceived susceptibility and health anxiety while simultaneously reducing objective risk for COVID-19. PPE use and social distancing would thus be negatively reinforced (via anxiety reduction) and adaptive by reducing objective risk.

Trait mindfulness has been described as a set of attributes where cognitive, affective, and physiological experiences are acknowledged and accepted from a nonreactive and nonjudgmental perspective (Shapiro, et al., 2006). From a functional contextual perspective, higher levels of trait mindfulness may allow an individual to more effectively evaluate health threat information, manage health anxiety, and cope with the emotional or physical discomfort that can arise from engaging in health protective behaviors (e.g., mask wearing, distancing). Higher levels of trait mindfulness should thus be associated with higher engagement in health protective behavior.

Research examining relationships between mindfulness-related variables and health protective behavior during the COVID-19 pandemic is scarce. Our exhaustive search of the literature and yielded only three published studies. All three measured mindfulness and health protective behaviors at a single time point and used cross-sectional analyses. Grover and colleagues (2020) collected data on a sample of 315 persons living in India from May 12, 2020 to May 17, 2020. Mindfulness was assessed with the short version of Mindfulness Attention and Awareness Scale (van Dam et al., 2010). Their results revealed that mindfulness was significantly associated with self-reported physical distancing. The effect size was substantial (*B* = .46, *p* < .001).

Haliwa and colleagues (2020) conducted a study using a sample of 374 MTurk workers, mindfulness (as measured by the Cognitive and Affective Mindfulness Scale – Revised). Regression analyses were used to predict health protective behaviors. Demographic, personality, perceived risk, objective risk, and mindfulness variables were predictors. Results indicated that mindfulness was a significant predictor of social distancing (*b* = .17, *p* = .02.).

O’Brien and colleagues (2021) conducted a cross-sectional analysis of 450 USA MTurk workers in April 2020. They found that perceived susceptibility and mindfulness related variables (as measured by the Five Factor Mindfulness Scale) predicted PPE use in regression analyses controlling for COVID-19 relevant demographic variables. Higher levels of FFMQ nonreactivity were associated with higher levels of PPE use. Interestingly, higher levels of FFMQ nonjudgment were associated with less PPE use.

The inverse relationship between FFMQ nonjudgement and PPE use was interpreted within the framework of the Unified Flexibility Model which synthesized the facets of all major mindfulness measure into a comprehensive functional contextual model (Rolffs, Rogge, & Daks, 2018). In the Unified Flexibility Model, the FFMQ nonjudgment subscale which is usually reverse scored (i.e., *Disagreement* with a statement such as “I make judgments about whether my thoughts are good or bad” is interpreted as an index mindfulness) is <u>not</u> reverse scored. The non-reversed scored scale (i.e., level of agreement with a statement such as “I make judgements about whether my thoughts are good or bad.”) is used as a measure “non-mindfulness” or psychological inflexibility – which in this case is a person’s tendency to judge thoughts and feelings. From this perspective, the relationship between the FFMQ nonjudgment scale and PPE use becomes more interpretable: Persons who *agreed* with statements indicating that they judged thoughts and feelings to be good/bad and right/wrong, also reported higher levels of adherence to COVID-19 PPE recommendations which they likely perceived to be “right” and “good.”.

### Summary

There are only three investigations of the relationship between mindfulness related constructs and COVID-19 health protective behavior. Two studies found a positive relationship between a general measure of mindfulness and social distancing. The other study found a positive relationship between FFMQ mindful nonreactivity and PPE use as well as a negative relationship between FFMQ nonjudgment and PPE use. The three studies used single time point measurement and cross-sectional analyses which limited causal inference. Covariation combined with temporal order are the sine qua non for causal inference (Haynes & O’Brien, 2000). The cross-sectional analyses of the three studies demonstrated covariation but not temporal precedence. Therefore, the inference that mindfulness *predicts* COVID-19 related PPE use and social distancing is limited.

In this investigation, we measured perceived susceptibility to COVID-19, mindfulness, and COVID-19 health protective behaviors at two time points separated by a 2-month interval during the early phases of the COVID-19 pandemic in the USA (April 2020 and June 2020). This allowed us to establish temporal order and evaluate the extent to which mindfulness-related variables predicted COVID-19 health protective behaviors over and above the many contextual factors (social, political, and individual circumstances) that were happening at the time.

## Methods

### Participants

This project was approved by the USA university Institutional Review Board on March 30, 2020. Amazon Mechanical Turk Workers (MTurk) were enrolled via CloudResearch from April 9, 2020 through April 18, 2020 (Litman, Robinson, & Abberbock, 2016). All participants provided informed consent. In the first wave of the study (Time 1), 450 participants were recruited (O’Brien et al., 2021). This number was sufficient for conducting suitably powerful regression analyses of up to 10 predictor variables (Wilson & Morgan, 2007). A follow-up survey announcement and link were posted on MTurk/CloudResearch on June 19, 2020. The link was kept active for four weeks.

A total of 178 MTurk workers completed the second survey (Time 2). Time 1 and time 2 surveys were matched using unique ID codes generated by participants at time 1, IP addresses, and demographic characteristics. Out of the 178 responses, we were able to unambiguously match 151. The demographic characteristics of these 151 participants are summarized in Table 1. The data used for this project can be accessed on the Harvard Dataverse at doi:10.7910/DVN/LIDGNS. This sample size was sufficient for evaluating up to 10 predictors in a regression analyses with adequate power.

**Table 1.**
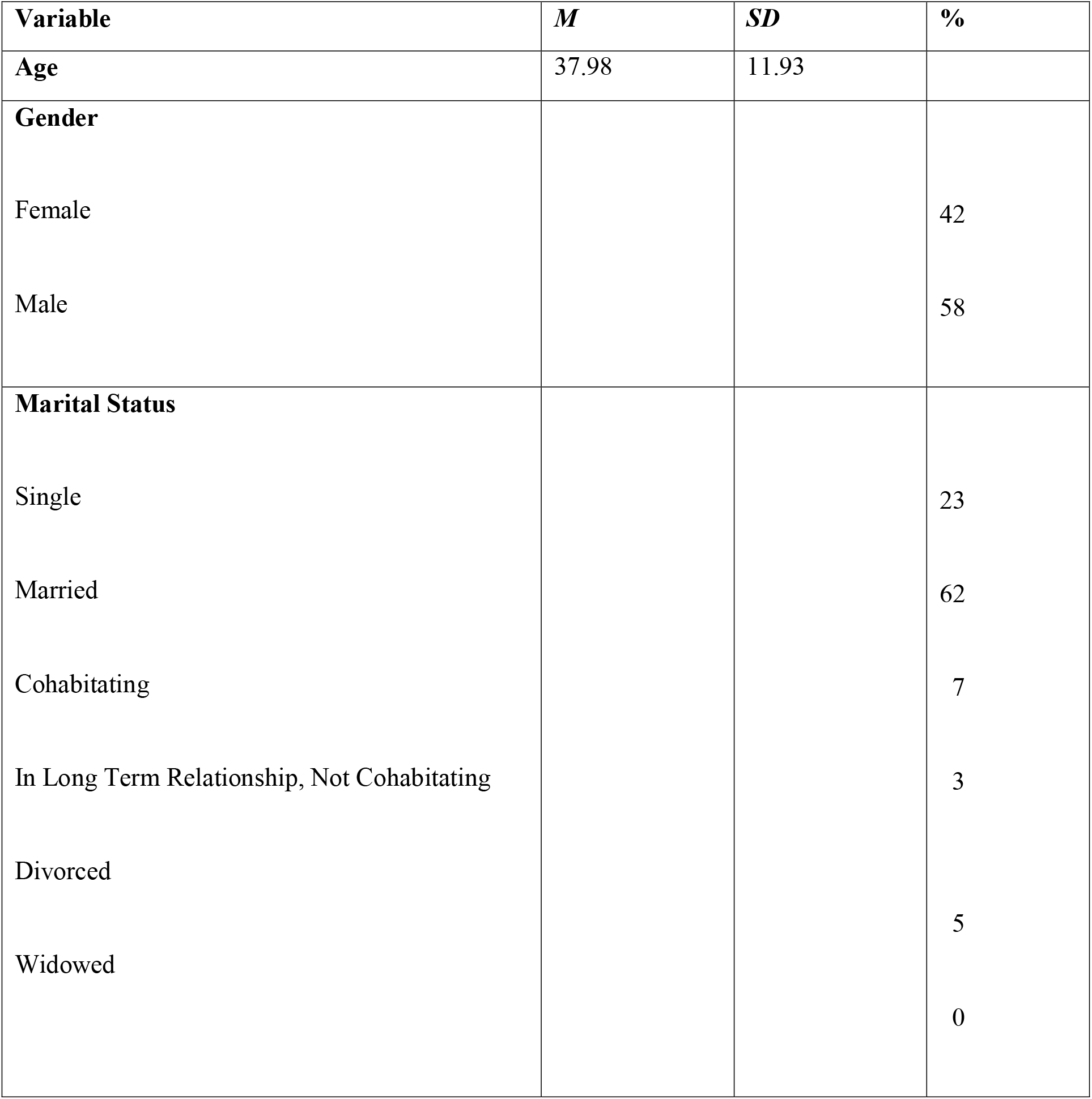
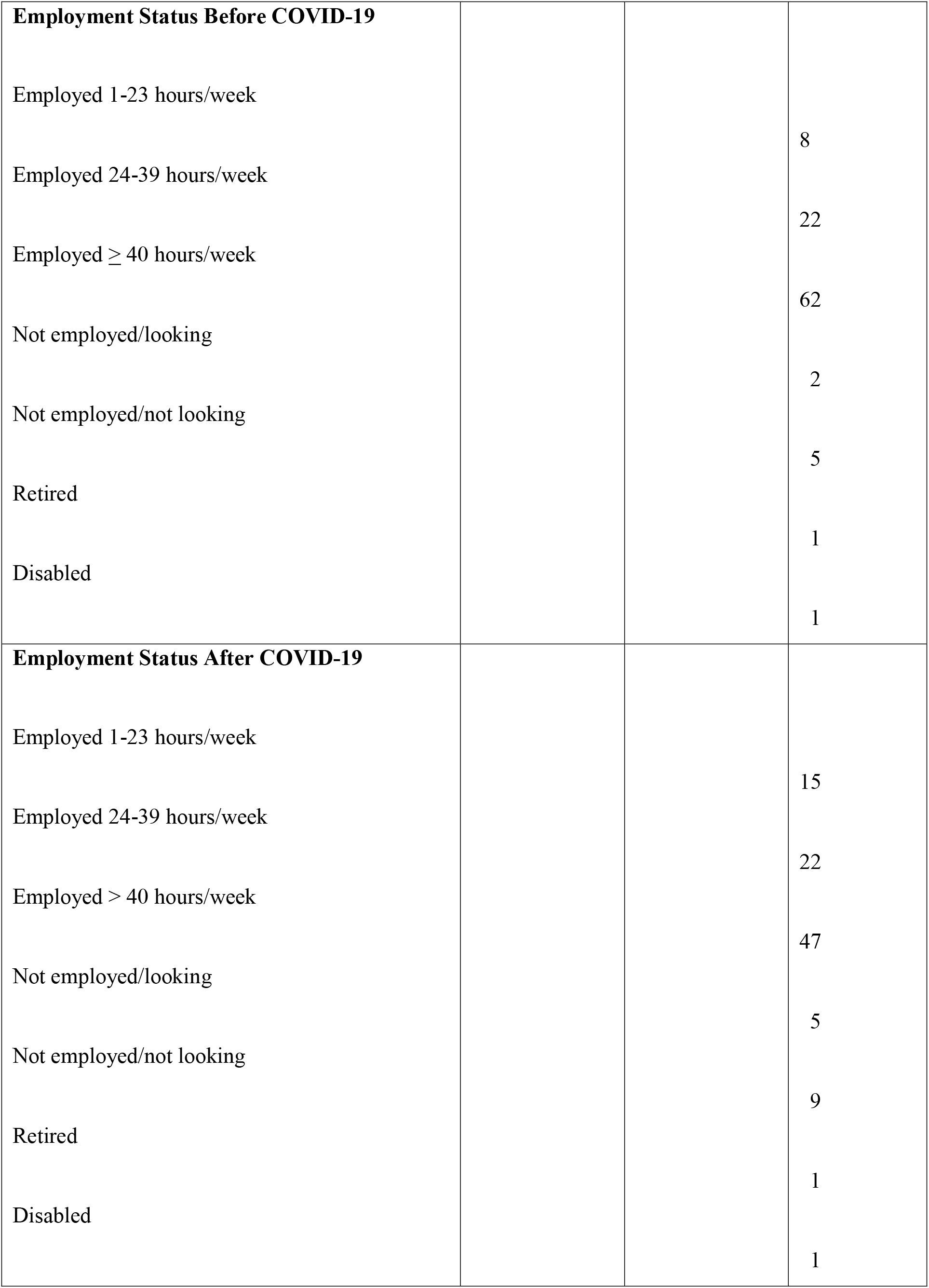
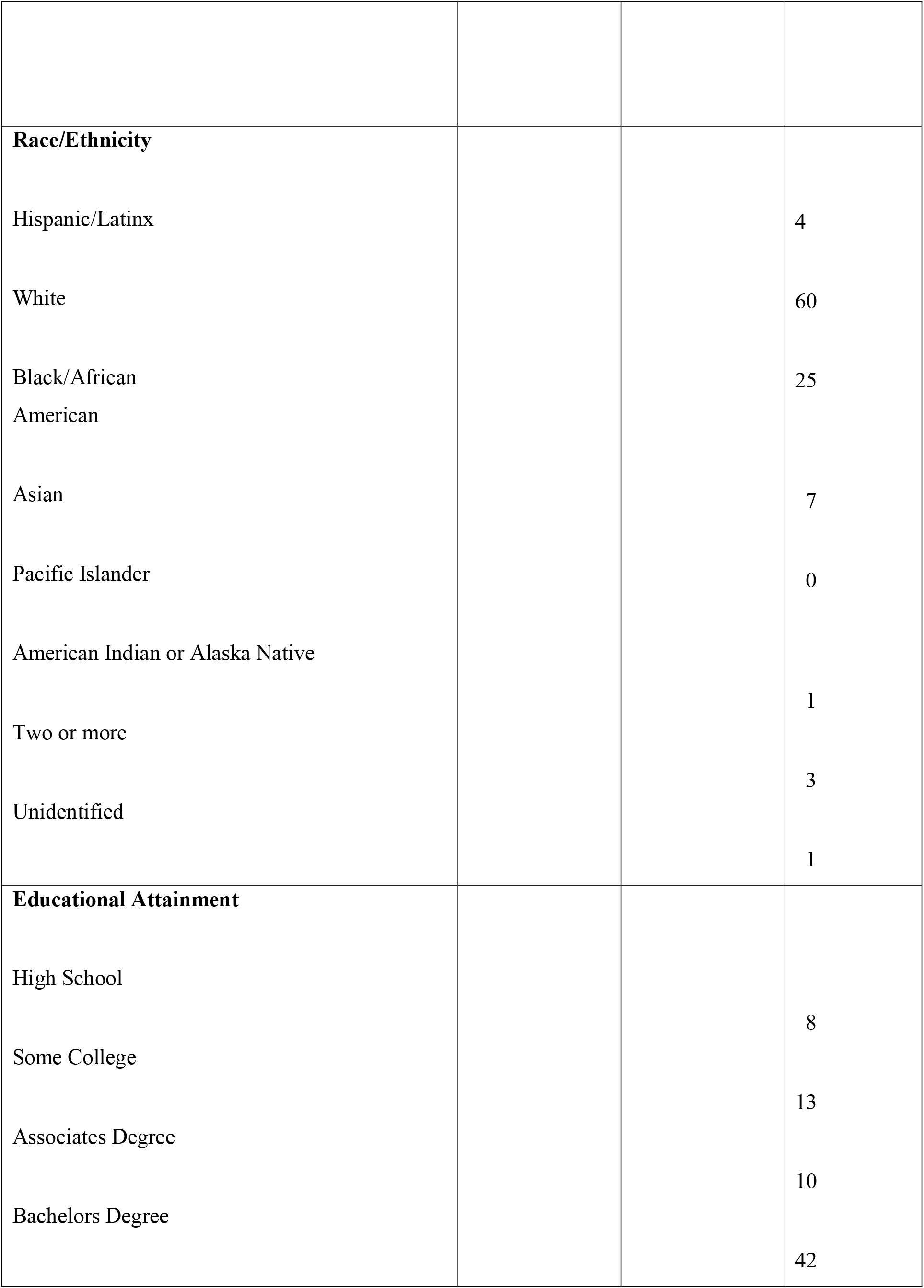
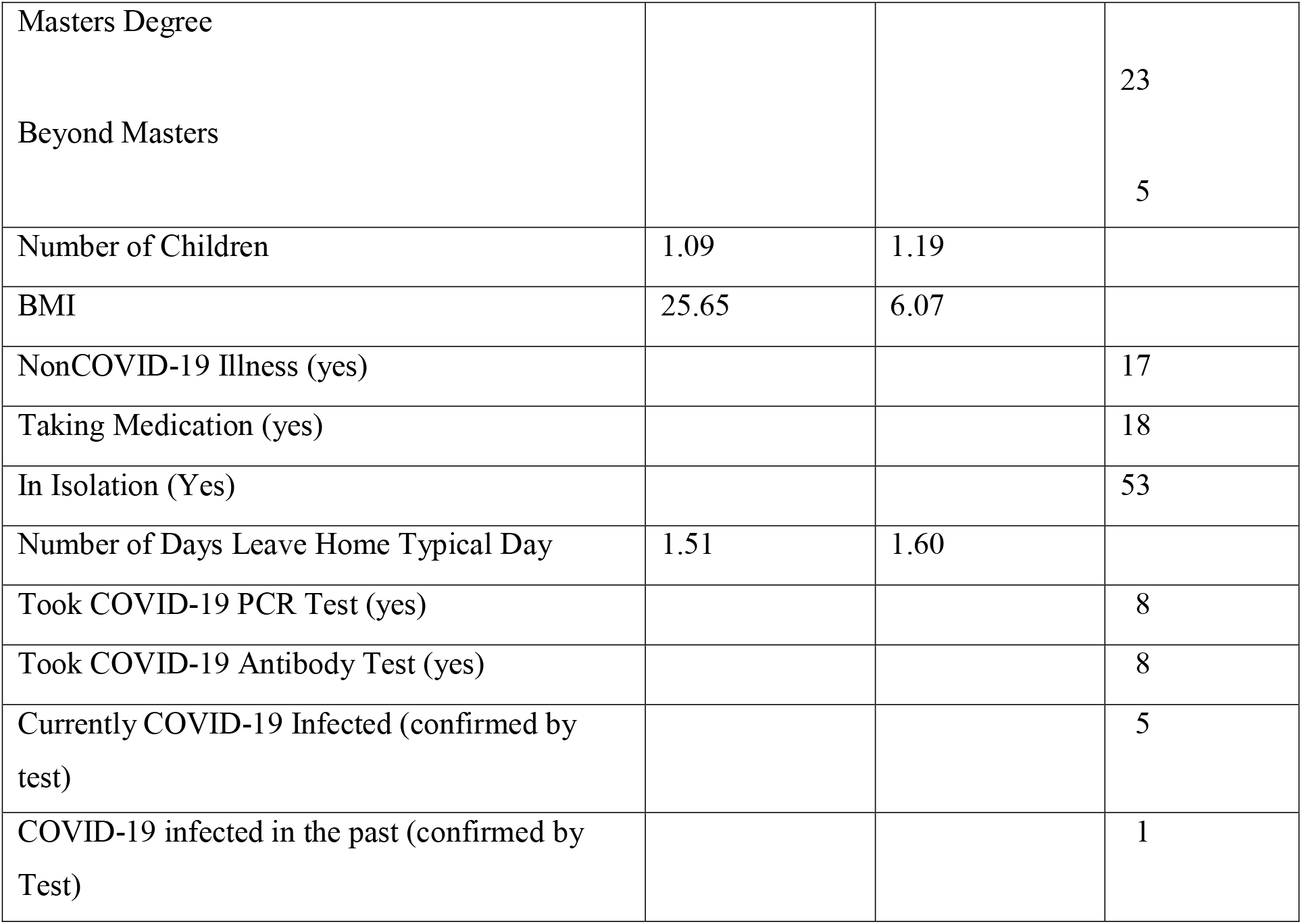
Demographic characteristics of Sample.

Although there are criticisms about the use of MTurk samples, *during the early stages of the COVID-19 pandemic MTurk provided one of the most workable systems for gathering data about COVID-19*. Several strategies were used to maximize the integrity of MTurk recruitment and data quality. First, the announcement only went to MTurk workers who had a high quality ratings. Second, we restricted participation to MTurk workers in the USA to avoid MTurk “farms.”. Third, the survey had 3 attention check items that could detect bots and uncareful rapid responding. Fourth, we added 3 captcha items that could detect bots. Fifth, all surveys were carefully examined for duplicate IP addresses, completion times, and data quality. Sixth, we added open ended questions. These open-ended questions were examined for evidence of bots. Bots tend to generate nonsensical responses or responses that copy and paste words from the open-ended question.

## Measures

### Demographics and COVID-19 Risk Factors

Information about demographic characteristics, employment, and living arrangements were collected. Participants also reported whether they were experiencing any non COVID-19 illnesses and listed medications they were taking as well as height and weight to calculate BMI. A composite risk factor score was generated by dichotomizing and summing across established risk factors (CDC, 2020; Millet et al, 2020): age (< 60 = 0, ≥ 60 = 1), illness (non COVID-19 risk factor illnesses = 0, established COVID-19 risk factor illnesses such as cardiovascular disease, diabetes, autoimmune disorders = 1), BMI (< 30 = 0, ≥ 30 = 1), race (African American = 1, all other = 0), sex (female = 0, male = 1), and occupation (healthcare = 1, all others = 0). The composite risk factor score could range from 0 to 6. The demographic and health questionnaire was used in previous mindfulness and health investigations.

### Perceived susceptibility to COVID-19

To assess perceived vulnerability to COVID-19 participants were asked to rate the likelihood of contracting COVID-19. Response options ranged from “no chance” to “certain” using a five-point scale.

### Mindfulness

The Five Facet Mindfulness Questionnaire-24 (FFMQ) was used to measure trait mindfulness (Baer, et al., 2006). The FFMQ-24 study has awareness, observe, describe, nonjudgment, and nonreactivity subscales. Using a larger MTurk sample collected at Time 1, O’Brien and colleagues (2020) reported the internal consistency for the entire FFMQ-24 was Cronbach’s alpha = .79 and the internal consistencies of each subscale were: awareness (α = .87), observe (α = .72), describe (α = .61), nonjudge (α = .80), and nonreactivity (α = .80). Higher scores on each subscale indicated higher levels of mindfulness.

### Health Protective Behaviors

The Preventive actions taken scale (PATS) was developed in late January 2020 in China as the pandemic was emerging (O’Brien, et al., 2020). The original 12-item measure assessed PPE use (masks, gloves), obtaining cleaning and preventive health supplies, avoiding contact with other people, avoiding travel, and avoiding risky foods. O’Brien and colleagues (2020) modified the PATS based on item analysis and factor analysis which resulted in an 8-item survey that assessed two domains of COVID-19 preventive health behavior: *PPE use and social distancing*. The internal consistencies of the two subscales were good (PPE Cronbach’s α = .77; social distancing Cronbach’s α = .76). Higher scores indicated more engagement in health protective behavior.

### Procedure

At Time 1 (April 9, 2020) an announcement was placed on Amazon Mechanical Turk via CloudResearch. The announcement read “The COVID-19 situation is creating worldwide challenges. In this survey study, university researchers hope to gain important useful information about how people are reacting to COVID-19 and coping with COVID-19.” Participation was limited to individuals who were at least 18 years old and residing in the United States. Participants received $1.00 for completing the survey. After providing consent participants were linked to the survey. A follow-up announcement was sent to every participant who completed the Time 1 survey on June 19, 2020. To match Time 1 and Time 2 surveys, participants were asked to generate a unique ID that they reported in each survey.

## Results

### Perceived Susceptibility to COVID-19 at Time 1 and Time 2

The difference in average perceived susceptibility at Time 1 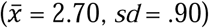 and Time 2 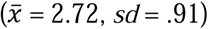 was nonsignificant. At both points of measurement, nearly 60% of the participants perceived themselves to have a 50/50 chance or higher risk for contracting COVID-19.

### Health Protective Behaviors at Time 1 and Time 2

A Time 1 versus Time 2 repeated measures MANOVA was conducted using the PATS subscales PPE use and social distancing as two dependent variables. A significant MANOVA main effect with a large effect size was observed for Time [Wilkes Lambda = .87; *F* (2, 142) = 10.95, *p* < .001; *η*^*2*^ = .13). The follow-up ANOVA for PPE use was significant [*F* (1, 142) = 9.34, *p* = .003; *η*^*2*^ = .06] and the follow-up ANOVA for social distancing was also significant [*F* (1, 142) = 5.17, *p* = .024; *η*^*2*^ = .06]. Both effect sizes fell into the medium classification.

The change from Time 1 to Time 2 for the two preventive behaviors were in opposite directions. PPE use increased from Time 1 to Time 2 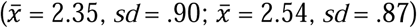 whereas social distancing declined from Time 1 to Time 2 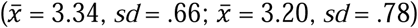.

### Correlations among Health Protective Behaviors, Risk Factors, Perceived Susceptibility, and Mindfulness

The intercorrelations among all predictor and dependent variables are presented in Table 2. The two PATS subscales were minimally correlated with each other suggesting that COVID-19 the two types of health protective behaviors are functionally distinct. Education and FFMQ nonreactivity were significantly associated with higher levels of PPE use while higher levels of FFMQ nonjudgment, awareness, and describe were associated with lower levels of PPE use.

**Table 2.**
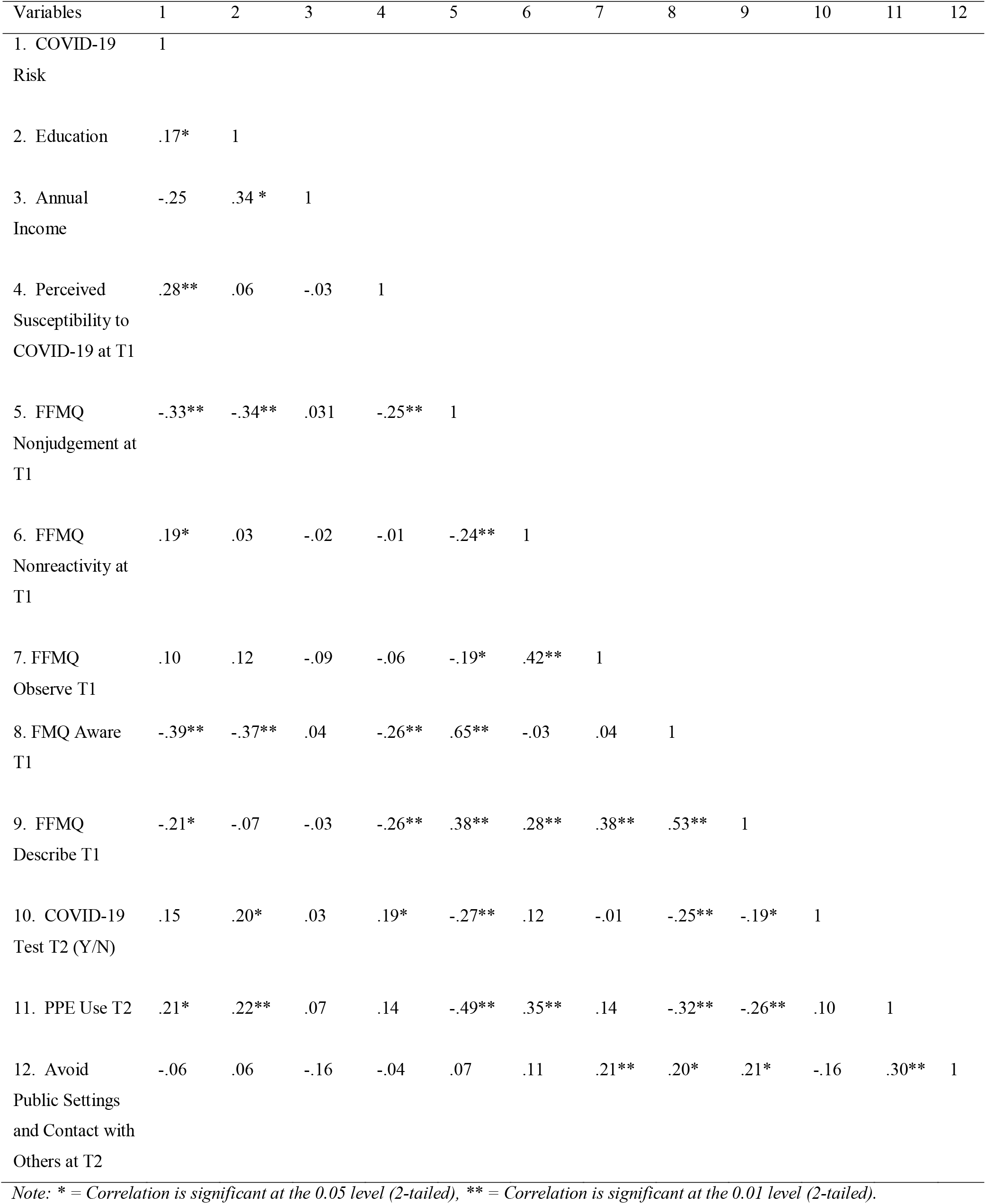
Correlations Among Predictors and Dependent Measures

### Predicting COVID-19 PPE Use Across a Two-month Interval

A three-step hierarchical regression was used to predict COVID-19 PPE use. <u>All of the predictor variables were from Time 1</u>. Step 1 predictor variables were the COVID-19 risk factor composite, educational attainment, and annual income. The step 2 predictor was perceived susceptibility to COVID-19. The step 3 predictor variables were the five FFMQ-24 subscale scores (aware, describe, observe, nonreactivity, nonjudgment). PPE use was measured at Time 2.

The first step of the regression was significant *F* (3, 140) = 6.04, *p* = .001; *R*^*2*^ = .12]. Step 2 did not produce a significant change in *R*^*2*^ [*F* (3, 140) = 1.58, *p* = .211; *R*^*2*^ change = .01]. Step 3 yielded a large and significant increase in *R*^*2*^ [*F* (3, 140) = 10.24, *p* < .001; *R*^*2*^ change = .24]. Therefore, the full regression model was interpreted. An analysis of individual standardized betas indicated that the COVID-19 risk factor composite was a significant predictor of PPE use (β = .15, *p* = .038). FFMQ-24 nonreactivity was significantly and positively associated with PPE use (β = .32, *p* < .001). FFMQ-24 nonjudgement and describe were inversely associated with PPE use (respectively: β = -.31, *p* = .002; β = - 25, *p* = .012).

### Predicting COVID-19 Social Distancing Across a Two-Month Interval

A three-step hierarchical regression was used to predict avoiding the PATS avoiding the public and contact with people subscale. The first step of the regression was nonsignificant *F* (3, 140) = 1.92, *p* = .129; R^*2*^ = .040]. Step 2 yielded a nonsignificant change in R^2^ [*F* (3, 140) = .816, *p* = .35; R^*2*^ change =.006]. Step 3 yielded a significant increase in R^2^ [*F* (3, 140) = 3.16, *p* = .010.; R^*2*^ change = .10]. Therefore, the full regression model was interpreted. An analysis of individual standardized betas indicated that education was significantly positively associated with Time 2 social distancing (β = .21, *p* = .031) while income was significantly negatively associated with social distancing (β = -.23, *p* = .012). FFMQ-24 awareness was significantly and positively associated with social distancing (β = .29, *p* = .014).

## Discussion

Across a two-month interval different aspects of mindfulness were significant predictors of PPE after controlling for objective risk, demographics, and perceived susceptibility. FFMQ nonreactivity was positively associated with PPE use. This finding was consistent with O’Brien and colleagues’ cross-sectional analyses which demonstrated that FFMQ nonreactivity was associated with PPE use at a single point in time (O’Brien et al., 2021). FFMQ nonjudgement and FFMQ describe were negatively associated with PPE use. The magnitudes of these relationships were substantial and two to three times larger than the average relationship between mindfulness variables and other nonCOVID-19 health behaviors reported by Sala, Rochfort, Lui, and Baldrum (2019) in their comprehensive meta-analysis of 125 studies. It is noteworthy that the magnitude of relationship across a two-month time interval was this substantial – particularly given the presence of many other COVID-19 influencing contextual factors (regional regulations, political messaging, social messaging, etc.).

There are several mechanisms through which FFMQ nonreactivity could promote higher PPE. First, it could be that people who have higher levels of nonreactivity have a history of being more effective in managing distressing thoughts and emotions. Applied to PPE use, this could take the form of being less apt to view PPE use as burdensome, distressing, risky, or undesired. When weighing the benefits and costs of PPE use these persons may be more inclined to engage in the PPE behavior given that there are fewer and/or less intense experienced costs (negative emotional reactions). O’Brien and colleagues (2019a) observed a similar finding when studying the relationship between mindful awareness and safety behavior among nursing aides. Nursing aides who reported higher levels of mindful awareness also reported higher levels of safety compliance. Furthermore, O’Brien and colleagues (2019b) found that training in Acceptance and Commitment Therapy (which has a significant mindfulness component) was associated with fewer days of work missed due to occupational injuries.

Second, it may be that mindful nonreactivity promotes better problem solving. A calm, non-distraught, and non-reactive stance would allow for a better evaluation of risks, benefits, and options for self-care via reduced psychophysiological activation. Multiple researchers have demonstrated that mindfulness is associated with improved creativity and problem solving (e.g., Ostafin & Kassman, 2012) as well as reduced levels of psychophysiological activation (Pascoe et al., 2017).

Third, it may be that higher mindful nonreactivity promotes enhanced self and other compassion. With the diminished psychophysiological reactivity and enhanced problem solving a person may be more aware of the need to care for self and others. These connections between mindfulness and enhanced compassion have been well-articulated among researchers exploring East-West integration of mindfulness theories and phenomena (Jarukasemthawee et al., 2019; Neff et al., 2008). Researchers exploring the neurovisceral integration and polyvagal theories have also documented biobehavioral pathways linking mindfulness, problem solving, and compassion (Ashar et al., 2016; Blair & Peters, 2003; Desbordes et al., 2012; Kim et al., 2020; Lutz et al., 2009; Thayer et al., 2009; Thayer & Sternberg, 2006; Wadden et al., 2018).

The negative relationship between PPE use and FFMQ nonjudgment was consistent with the prior cross-sectional analyses by O’Brien and colleagues (2021). This finding indicates that participants who more strongly *agreed* that thoughts, feelings, and actions were “right or wrong” also reported that higher PPE use. It may be that these persons also viewed PPE use as “right.” Thus, the relationship between the two measures may be driven by values-guided behavior.

There are limitations in this study that merit discussion. MTurk workers may have characteristics that differ from other samples of individuals. Chandler and Shapiro (2016) reported that MTurk samples can be more representative of the USA population relative to convenience samples but less representative than probability samples used in epidemiological research. However, Chandler and Shapiro (2016) also pointed out that national probability can be biased because they rely on telephone methods that can over sample older and more conservative participants. There are some strengths to MTurk samples. During the COVID-19 lockdown, online data gathering was the *only* way to gather complex survey data from diverse populations from broad geographic locations. Additionally, we adopted “best practices” by screening for response quality, using attention checks, and making sure that signaling cues were not in the surveys (Ophir, et al., 2019; Cheung et al., 2017).

A total of 450 persons completed the Time 1 survey and 151 (34%) completed the Time 2 survey. The 34% completion rate for this study is consistent with other MTurk research. Auer et al., (2021) conducted a study examining retention levels for MTurk workers across a 30-day interval using a 30-minute survey (which is similar in length to the survey used in this study). With $1.00 compensation for completing each survey (which is equal to our payments), they observed a 35% retention rate.

We argue that mindfulness may promote less psychophysiological reactivity, better problem solving, and improved compassion. We did not have measures of these constructs. In future investigations it would be beneficial to examine the extent to which persons scoring higher on trait mindfulness demonstrate lower levels of psychophysiological activation with mask wearing and other health protective behaviors. Additionally, it would be quite illuminating to evaluate the extent to which PPE use is driven not only by self-oriented perceptions of susceptibility, but also by compassion towards others.

## Conclusion

These mindfulness results highlight complex multidimensional nature of mindfulness and its relationships with COVID-19 health protective behaviors. Promoting mindfulness skills focused on nonreactivity may be able to improve adherence and improved COVID-19 health protective behaviors. Mindfulness training has been shown to promote improved health protective behavior among healthcare professionals, better problem solving, lower psychophysiological reactivity to stress, and improved compassion. It is reasonable to conclude that mindfulness training could also be important for promoting COVID-19 health protective behavior.

A public-health level mindfulness promotion would augment the standard COVID-19 health messaging (which typically emphasizes perceived susceptibility and efficacy of health protective behaviors) with information about how to manage COVID-19 related anxiety, mindful management of PPE discomfort, and mindful self and other compassion. The promotion could occur via online messaging, educational interventions, and workplace-based mindfulness training.

## Compliance with Ethical Standards

This project was reviewed and approved by the Bowling Green State University Institutional Review Board (project #1562479-4). All participants were provided with a consent document. All participants provided informed consent prior to taking the surveys.

## Data Availability

The data used for this project can be accessed on the Harvard Dataverse at doi:10.7910/DVN/LIDGNS.

doi:10.7910/DVN/LIDGNS

